# Safety and cognitive pharmacodynamics following dose escalations with 3-methylmethcathinone (3-MMC): a first in human, designer drug study

**DOI:** 10.1101/2024.11.12.24317168

**Authors:** JG Ramaekers, JT Reckweg, NL Mason, KPC Kuypers, SW Toennes, EL Theunissen

**Affiliations:** Faculty of Psychology and Neuroscience, Maastricht University, Maastricht, The Netherlands; Goethe University, Institute of Legal Medicine, Frankfurt, Germany

## Abstract

3-Methylmethcathinone (3-MMC) is a designer drug that belongs to the group of synthetic cathinones. The compound has been scheduled in many jurisdictions because of public health concerns associated with excessive use. To date, there are no clinical studies that have evaluated the risk profile of 3-MMC in the recreational range of low to moderate doses. The current, first-in-human study (N=14) assessed the impact of three escalating doses of 3-MMC (25, 50 and 100 mg) on vital signs, neurocognitive function, state of consciousness, appetite and drug desire, in a cross-over, placebo-controlled trial. A battery of neurocognitive tests and questionnaires as well as measures of vital signs were repeatedly administered up to 5 hours after dosing. Overall, 3-MMC caused dose-dependent increases in heart rate and blood pressure, though not of clinical significance, and feelings of subjective high. Additionally, 3-MMC induced dose-related enhancement of task performance across several neurocognitive domains, including processing speed, cognitive flexibility, psychomotor function, attention and memory. Impulse control was not affected by 3-MMC. Participants also reported mild increases in dissociative and psychedelic effects, decreased appetite, and gave greater ratings of liking and wanting for 3-MMC that were transient over time. Overall, the cardiovascular, psychostimulant and psychotomimetic profile of 3-MMC appears consistent with that of compounds structurally related to amphetamine. It is concluded that low to moderate doses of 3-MMC were well tolerated and safe and that potential health risks might only occur at high or excessive doses of 3-MMC.

## Introduction

3-Methylmethcathinone (3-MMC) is a synthetic analogue of the natural psychoactive substance cathinone, a psychostimulant present in the leaves of the khat shrub [1]. Derivatives of cathinones such as 3,4-methylenedioxy-N-methylcathinone (methylone), 4-methylmethcathinone (4-MMC, mephedrone) or 3,4-methylenedioxypyrovalerone (MDPV) have come to prominence as designer drugs or ‘legal highs’ and have been the subject of critical review by public health organizations [2–5], resulting in restrictive legislation in several countries [6,7].

Cathinones are structurally related to amphetamines, which is why there used to be an interest in the 1940s and 50’s to synthesize cathinones as medicinal products to suppress appetite, reduce symptoms of depression or stimulate brain activity [1]. Likewise, the capacity of synthetic cathinones such as 3-MMC to increase alertness, wakefulness, euphoria and to suppress appetite has been a main driver of recreational use [1,2,8]. 3-MMC is a monoamine transporter substrate that potently inhibits noradrenaline, dopamine and serotonin reuptake, but in addition also displays strong binding affinities for adrenergic receptors [9], which are known to modulate stimulant-induced behavior [10]. Moreover, 3-MMC also shows binding affinities for 5HT1A and 5HT2A receptors [9], but the role of these receptors to the psychoactive effects of 3-MMC, either through stimulation or antagonism, is currently unknown. Users describe the effects of 3-MMC as similar to, but less potent and less intense than those of 3,4-methylenedioxymethamphetamine (MDMA) and 4-MMC [11]. Besides psychostimulant effects, 3-MMC may also increase feelings of empathy and affection and cause sensory enhancement [12,13].

Synthetic cathinones are administered recreationally by nasal insufflation (snorting), orally, and by intravenous injection [13]. For 3-MMC, oral doses range from 25–75 mg (light), 75-150 (common) and 150–300 mg (high), although the use of even higher doses has been reported as well [4,14,15]. 3-MMC administrations are sometimes repeated to prolong their desired effects resulting in total doses as large as 2 grams during a single session [11]. Estimates of ‘typical’ doses are however based on self-reports, and should be interpreted with caution, as the users are not always aware of the actual substance or amount used.

Besides their desired psychostimulant effects, synthetic cathinones may also cause serious adverse events, including acute sympathomimetic (e.g., blurred vision, dry mouth, hyperthermia, and mydriasis) and cardiovascular (e.g., increase in blood pressure and heart rate) changes. Additionally, there is a risk of psychological dependence, abuse potential, and withdrawal symptoms. [13]. Most knowledge on adverse events with synthetic cathinones comes from case-studies, often involving the use of high doses combined with alcohol or other drugs. In the case of acute 3-MMC intoxication, case reports on adverse events [4,5,16,17] have included fatigue, a reduced level of consciousness, aggression, agitation, psychomotor impairment and tachycardia. Less frequently reported adverse events included concentration difficulties, tingling in the arms and legs, diaphoresis, seizures, hyperthermia, and rhabdomyolysis. A small number of fatal intoxications have been associated with the use of 3-MMC, although in the majority of these cases other drugs were present as well [12,18].

To date, no clinical studies have been conducted with synthetic cathinones that could serve as a scientific base for risk evaluations, particularly at the low and moderate dose ranges that are most common among users. The current placebo-controlled, first-in-human study therefore aimed to determine the safety profile of low to moderate doses (25, 50, and100mg) of 3-MMC and monitored vital signs, psychomotor and cognitive function, state of consciousness, appetite and desire for 3-MMC for up to 5 hours after acute administration.

## Methods

### Participants

Fourteen participants (9 males, 5 females) entered the study, of whom 12 completed all treatment conditions. Two participants dropped out without completing all treatment conditions for reasons unrelated to the study. Participants’ age ranged between 19 and 35 years (mean (SD): 22.9 (4.3)). All participants had previous experience with stimulant drugs such as MDMA (11 participants), amphetamine (5 participants), cocaine (7 participants), or synthetic cathinones (4 participants). History of cannabis use, psychedelics, and ketamine was reported by 12, 9, and 4 participants respectively. All participants consumed alcohol. Participants were recruited through word-of-mouth and advertisements placed in Maastricht University buildings. All participants provided written informed consent and underwent a medical examination including a physical examination, routine laboratory tests (e.g., clinical chemistry, hematology, serology, urinalyses) vital signs and electrocardiogram (ECG). Inclusion criteria included: (a) age 18–40 years (b) absence of any major medical, endocrine, and neurological condition (c) free from psychotropic medication (d) good physical health (e) body mass index within 18.5-28 kg/m2 and (f) experience with psychostimulants in the past 12 months. Exclusion criteria included: (a) addiction (b) history of psychiatric or neurological disorder (c) pregnancy or lactating (d) cardiovascular, gastrointestinal, hepatic or renal abnormalities (e) excessive smoking (>15 cigarettes per day) or drinking (>20 standard units of alcohol per week) (f) hypertension (diastolic > 90 mmHg; systolic > 140 mmHg), (g) serious side effects of previous use of psychostimulants and (h) being a blood donor.

### Design and treatments

This study was conducted according to a phase 1, single-blind, placebo-controlled, cross-over design. Participants received single doses of 3-MMC (25, 50 and 100) or placebo on separate days. 3-MMC was dissolved in approximately 150 mL of a bittering agent (bitter lemon) and administered orally. Placebo drinks consisted solely of bitter lemon. An escalating dosing scheme was used. Participants received one of the following treatment orders: 0–25-50-100mg, 25–0–50-100, 25-50-0-100mg or 25-50-100–0mg. Conditions were separated by a minimum washout period of 7 days to avoid carry-over effects. During each 3-MMC administration day, participants were closely monitored and under medical supervision.

An independent Safety Group (SSG) was installed to evaluate vital and behavioral data collected after each dose during the first batch of 6 participants. The 2nd batch of participants only started after a positive SSG evaluation of all adverse events experienced during the 1st batch. The study was conducted according to the code of ethics on human experimentation established by the declaration of Helsinki (1964) and its amendments, and was approved by the Medical Ethics Committee of Maastricht University. A permit from the Dutch drug enforcement administration was acquired for obtaining, storing, and administering 3-MMC. Participants received monetary compensation for their participation in the study. The study was registered with the Dutch Central Committee of Research involving Human Subjects (CCMO) under trial number: NL84174.068.23.

### Procedure

Participants were asked to refrain from drug use at least one week prior to the start of the study, and to refrain from drug use throughout the entire study. Participants were not allowed to consume alcohol (48h), caffeine-containing beverages (24h) or tobacco before and during experimental sessions and were requested to arrive well rested. An alcohol breathalyzer and urine drug screen were conducted on test days after arrival of the participants. Urine drug screens checked for the presence of amphetamine, barbiturates, benzodiazepines, cocaine, methamphetamine, morphine, methadone, phencyclidine and tetrahydrocannabinol (THC) Treatments were administered only after verifying negative results for drug and alcohol screens, and, in the case of female participants, after confirming negative pregnancy tests. Throughout the 5 hours following treatment, subjective drug effects, vital signs and neurocognitive function were repeatedly assessed under medical supervision. Table 1 provides a schedule of safety and pharmacodynamic measures collected on treatment days. Participants underwent a training session before the experimental sessions to become acquainted with the neurocognitive tests and reduce practice effects.

**Table 1.**
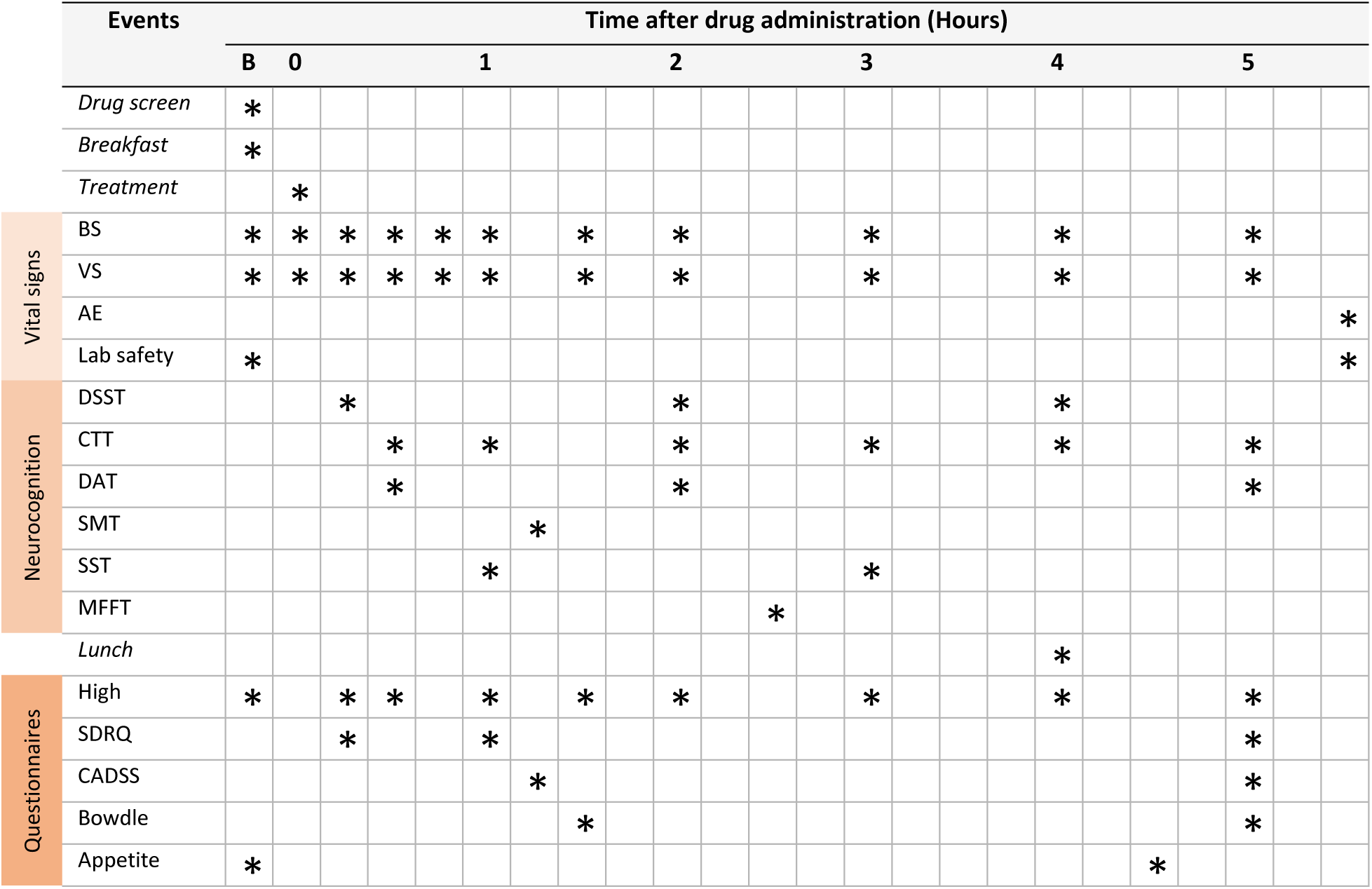
Schedule of safety and cognitive pharmacodynamic measures throughout a treatment day. BS=blood sample; VS= vital signs; AE=adverse events; DSST=digit symbol substitution task; CTT= critical Tracking Task; DAT= divided attention task; SMT= spatial memory task; SST=stop signal task; MFFT=matching familiar figures task; High=visual analogue scale (subjective high); CADSS=clinician administered dissociative states scales; SDRQ=sensitivity to drug reinforcement questionnaire; B=baseline.

### Vital signs and drug concentrations

Heart rate, blood pressure and temperature were monitored at regular intervals (Table 1). A lab safety blood and urine sample were taken at screening, baseline, and at the end of a test day for hematology, clinical chemistry and urinalysis. Blood samples to quantify 3-MMC concentrations were collected at baseline and at regular intervals after drug administration (see Table 1). Blood samples were centrifuged immediately and the serum was subsequently frozen at −30°C until analyses for pharmacokinetic assessment. 3-MMC serum concentrations were quantified by a forensic routine analysis method (liquid-liquid extraction of 0.2 ml serum, analysis using liquid chromatography-tandem mass spectrometry (LC-MS/MS), 1 ng/ml lower limit of quantification). Adverse events were documented at the end of treatment days. Participants were also asked to note any adverse events for 72 hours following dosing.

### Neurocognitive measures

A battery of tasks was included to assess psychomotor function, attention, decision-making, impulsivity and memory.

The Digit Symbol Substitution Task (DSST) [19] is a computerized version of the original paper and pencil test taken from the Wechsler Adult Intelligence Scale that measures processing speed, attention, and cognitive flexibility. In this task, participants are presented with an encoding scheme displayed as a row of squares at the top of the screen, where nine digits are randomly paired with specific symbols. The participant’s objective is to match each digit with its corresponding symbol from the encoding list and click the appropriate response button. The performance measure is the number of digits correctly encoded within a 3-minute time frame.

The Critical Tracking Task (CTT) [20] assesses a participant’s ability to manage a displayed error signal in a first-order compensatory tracking task. In this task, participants must make compensatory movements that increase in frequency over time. Eventually, the participant’s response frequency falls behind the error signal by 180°, causing their response to amplify the error rather than correct it, leading to a loss of control. The frequency at which this loss of control happens is known as the ‘critical frequency,’ or ‘lambda-c.’

The Divided Attention Task (DAT) evaluates a person’s ability to simultaneously manage two tasks [21]. The primary task involves using a joystick to continuously keep a cursor centered on a display by counteracting its horizontal movement. The tracking error is quantified as the absolute distance (in millimeters) between the cursor’s position and the center. For the secondary task, participants must monitor 24 single digits located at the corners of the computer screen and respond as quickly as possible by lifting their foot from a pedal whenever the target number ‘2’ appears. The dependent variables in this task are the mean tracking error and the mean reaction time to the targets.

The Spatial Memory Task (SMT) includes two phases: an immediate relocation phase and a delayed relocation phase. In the immediate relocation phase, participants complete six trials where they are shown a series of ten black-and-white pictures (totaling 60 pictures) at different locations on a computer screen. They must remember and indicate the locatio of each picture. After a 30-minute interval, the delayed relocation phase begins, during which the pictures reappear in random order at the center of the screen, and participants must again indicate the correct locations. The dependent variables measured in this task are the number of correct immediate relocations and the delayed relocation score [22].

The Stop Signal Task (SST) assesses motor impulsivity, defined as the inability to inhibit a pre-cued response, leading to errors of commission. Participants are required to respond quickly to visual go-signals and inhibit their response if a subsequent visual stop-signal, represented by an asterisk ("*"), appears in one of the four corners of the screen. The total duration of the task is approximately 8 minutes. The main dependent variables include the number of commission errors (failure to inhibit a response on no-go trials) and omission errors (failing to respond on go trials) [23].

The Matching Familiar Figures Test (MFFT) measures cognitive impulsivity. It involves showing a target figure on the left side of the screen and six alternative figures on the right, with only one matching the target exactly. Participants must identify the matching figure by pressing a corresponding keyboard number. Incorrect selections trigger a beep, prompting a retry. Each participant completes 2 practice trials followed by 20 test trials. Key metrics recorded are the mean latency of the first response and the total number of errors. These are used to calculate two scores after z-transformation: the Impulsivity score (I-score), which is the difference between the standardized scores of errors and standardized score of the mean latency to first response (Zerror - Zlatency), and the Efficiency score (E-score), which is the inverse of the sum of these standardized scores (1 - (Zerror + Zlatency)) [24].

### Subjective Questionnaires

Subjective drug experience (“high”) was measured using a 100 mm visual analogue scale with “not influenced by 3-MMC at all” at one end and “very influenced by 3-MMC” at the other end of the line.

The Sensitivity to Drug Reinforcement Questionnaire (SDRQ) asked two questions: How pleasant is using 3-MMC right now; How much do you want to use 3-MMC right now. Subjective valence of liking and wanting was scored on a 5-point scale: 1=somewhat; 2=slightly; 3=moderately; 4=very and 5=extremely.

The Clinician-Administered Dissociative States Scale (CADSS) [25] is a standardized measure of dissociative symptomatology. It comprises 19 subjective items, ranging from 0 ‘not at all’ to 4 ‘extremely. It is divided into 3 components: 1) depersonalization, 2) derealization and 3) amnesia. Summed together, these subscales form a total dissociative score.

The Bowdle Visual Analogue Scales (B-VAS) measures psychedelic effects using a 13-item visual analogue scale [26]. From these, log-transformed composite scores of ‘internal perception’ (i.e perception of the inner world) and ‘external perception’ (i.e. perception of the external world), as well as composite scores of ‘feeling high’ and ‘drowsiness’ are calculated.

Feelings of hunger and appetite were also rated by participants by means of 10 cm visual analogue scales after breakfast, lunch,and 10 hours after treatment (at home).

### Statistics

The effects of 3-MMC on all dependent measures were analyzed using a Linear Mixed Model with a restricted maximum likelihood method (REML). Model parameters included Dose, Time (after dosing) and Dose × Time as fixed effects, and a random intercept. The main effect of Dose was further segregated by pairwise comparisons between each 3-MMC dose and placebo (across time points). A first-order autoregressive residual covariance structure was used. The alpha criterion significance level was set at p = 0.05. All statistical tests were conducted with IBM SPSS version 27.0.

## Results

### Safety, subjective high and drug concentrations

Laboratory safety analyses, including hematology, clinical chemistry, and urinalysis, did not reveal clinically relevant deviations from normal ranges. Figure 1 presents the mean (SE) values for subjective experience, heart rate, systolic and diastolic blood pressure, and temperature following three doses of 3-MMC and placebo. LMM analyses showed main effects of Dose and a Dose by Time interaction respectively for subjective high (F_3,63.319_=17.440, p<.001; F_24,287.353_=7.407, p<.001), heart rate (F_3,67.452_=21.499, p<.001; F_30,338.168_=3.066, p<.001), systolic blood pressure (F_3,80.156_=29.620, p<.001; F_30, 341.160_=3.448, p<.001) and diastolic blood pressure (F_3,94.293_=24.338, p<.001; F _30,322.324_=2.413, p<.001). The main effect of Dose on temperature approached significance (F_3,90.448_=2.577, p=.059) and did not interact with Time. Drug-placebo comparisons showed that 3-MMC doses of 25mg (p=.045), 50mg (p<.001) and 100mg (p<.001) significantly increased subjective high. Drug-placebo comparisons furthermore revealed that 50 and 100mg 3-MMC increased systolic blood pressure (p=<.001, p<.001), diastolic blood pressure (p=.020, p<.001), and heart rate (p<.001, p<.001) respectively. Increments in temperature after a 100mg dose approached significance (p=.064).

**Figure 1.**
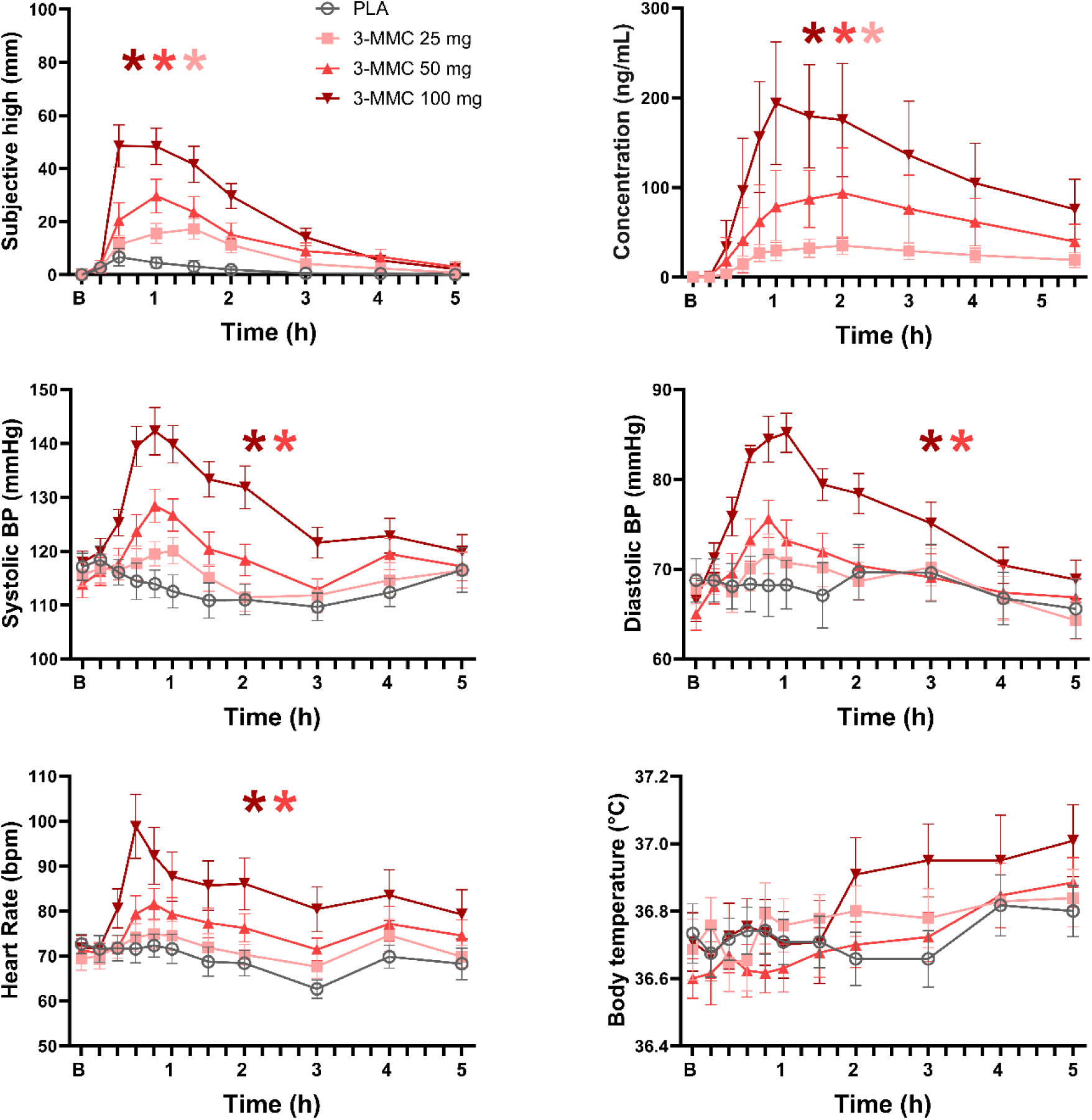
Mean (SE) ratings of subjective high and mean (SD) 3-MMC serum concentration as a function of time after dosing are shown in the upper row. The lower rows show mean (SE) vital signs (heart rate, systolic and diastolic blood pressure, temperature) throughout each treatment condition (B=baseline; A * denotes a significant (p < .05) contrast between 3-MMC and placebo across all time points, with the color of the * representing the 3-MMC dose level.

Mean (SD) concentrations of 3-MMC during a 5.5 hour time window are shown in Figure 1. 3-MMC concentrations varied with Dose (F_3, 46.439_=93.867,p<.001) and Dose by Time (F _30,321.699_=11.741, p<.001) after dosing. Drug-placebo contrasts showed dose related increments in 3-MMC concentrations after 25 (p<.001), 50 (p=.027), and 100 (p<.001) mg. Across all participants and all dose conditions the time to reach maximal 3-MMC concentrations (t_max_) varied between 0.75 and 3 (median 1.5) hours, while elimination half-life (t_½_) varied between 1.5 and 6.0 (median 3.0, interquartile range 2.5 - 3.6) hours. Tmax and t_½_ did not differ between doses.

### Neurocognition

Figure 2 presents the mean (SE) neurocognitive performances as a function of time after dosing in each treatment condition. LMM analyses showed main effects of Dose for the number of correct substitutions (F_3,53.054_=7.216, p<.001) in the DSST, lamda-c (F_3,55.870_=15.709, p<.001) in the CTT, tracking error (F_3,50.808_=22.123, p<.001) in the DAT, delayed relocations (F_3, 27.363_=4.083, p=.016) in the SMT, and efficiency in the MFFT (F_3, 22.884_=13.724, p<.001). There were no effects of Dose on measures of the SST. Dose by Time interactions did not reach significance for any of the neurocognitive measures. Drug-placebo contrasts revealed that 3-MMC increased the number of correct substitutions at 50mg (p=.005) and 100mg (p<.001), improved tracking performance (lamda-c) at 50mg (p<.001) and 100mg (p<.001), and reduced tracking error at 25mg (p<.001), 50mg (p<.001) and 100mg (p<.001), and increased the number of delayed relocations at 25mg (p=.007) and 50mg (p=.006). A dose of 50mg 3-MMC also increased efficiency in the MFFT as compared to placebo (p=.016).

**Figure 2.**
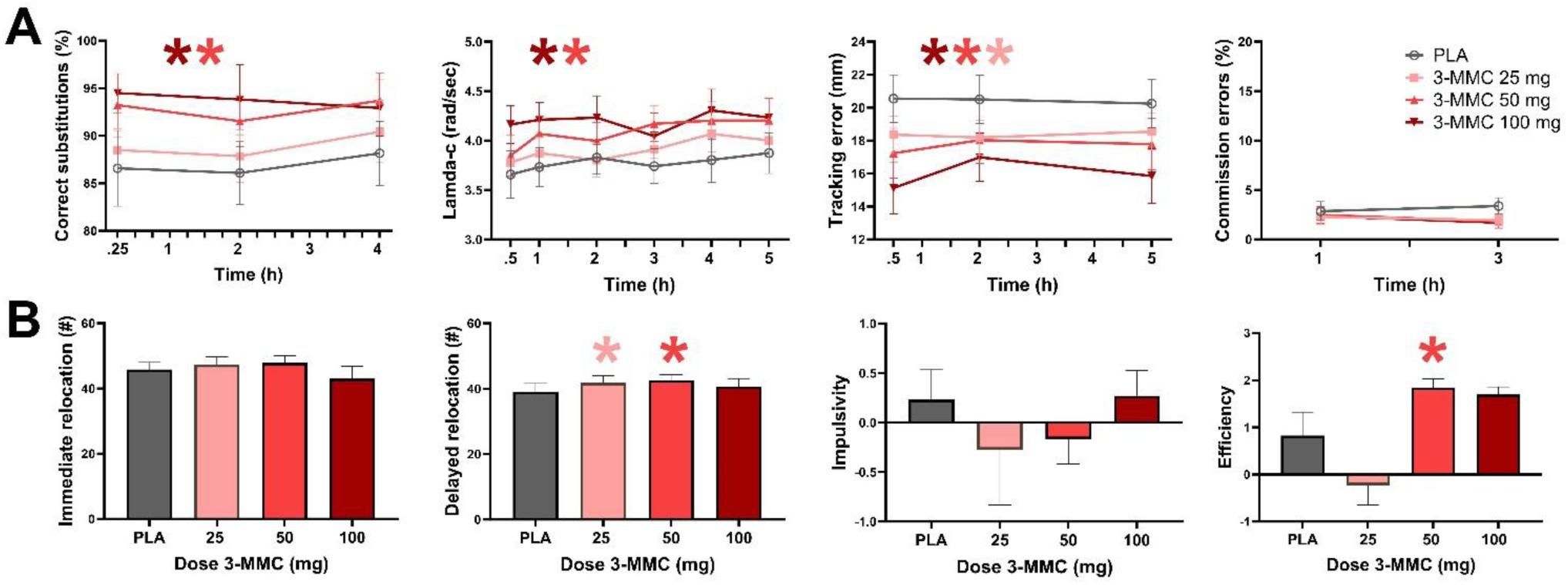
Panel A shows mean (SE) performances at the DSST (correct substitutions), CTT (lambda-c), DAT (tracking error) and SST (commission errors) as a function of time after dosing. Panel B shows mean (SE) performance at the SMT (immediate and delayed relocation) and MFFT (impulsivity, efficiency). A * denotes a significant (p < .05) contrast between 3-MMC and placebo across all time points, with the color of the * representing the 3-MMC dose level.

### Questionnaires

Figure 3 shows mean (SE) ratings of dissociative and psychedelic effects as assessed with the CADSS and Bowdle in every treatment condition. LMM analyses showed main effects of Dose on CADSS ratings of derealization (F_3,53.156_=7.218, p<.001) and total score (F_3,53.596_=6.012, p<.001), and on Bowdle ratings of high (F_3,54.128_=15.779, p<.001), internal (F_3,43.061_=5.472, p=.003) and external (F_3,39.601_=4.955, p=.005) perception. Dose by Time interactions were found for ratings of high (F_3,67.398_=8.316, p<.001) and external perception (F_3,69.695_=2.840, p=.044). Drug-placebo contrasts revealed increments in derealization at 25mg (p=.022), 50 mg (p<001) and 100mg (p<.001), increments in depersonalization at 100mg (p=.035) and increments in CADSS total score at 50mg (p=.003) and 100mg (p<.001). Drug-placebo contrasts also revealed increments in ratings of high at 25mg (p=.018), 50mg (p<.001) and 100mg (p<.001), increments in internal perception at 100mg (p<.001), and increments in external perception at 25mg (p=.006), 50mg (p=.005) and 100mg (p=.001).

**Figure 3.**
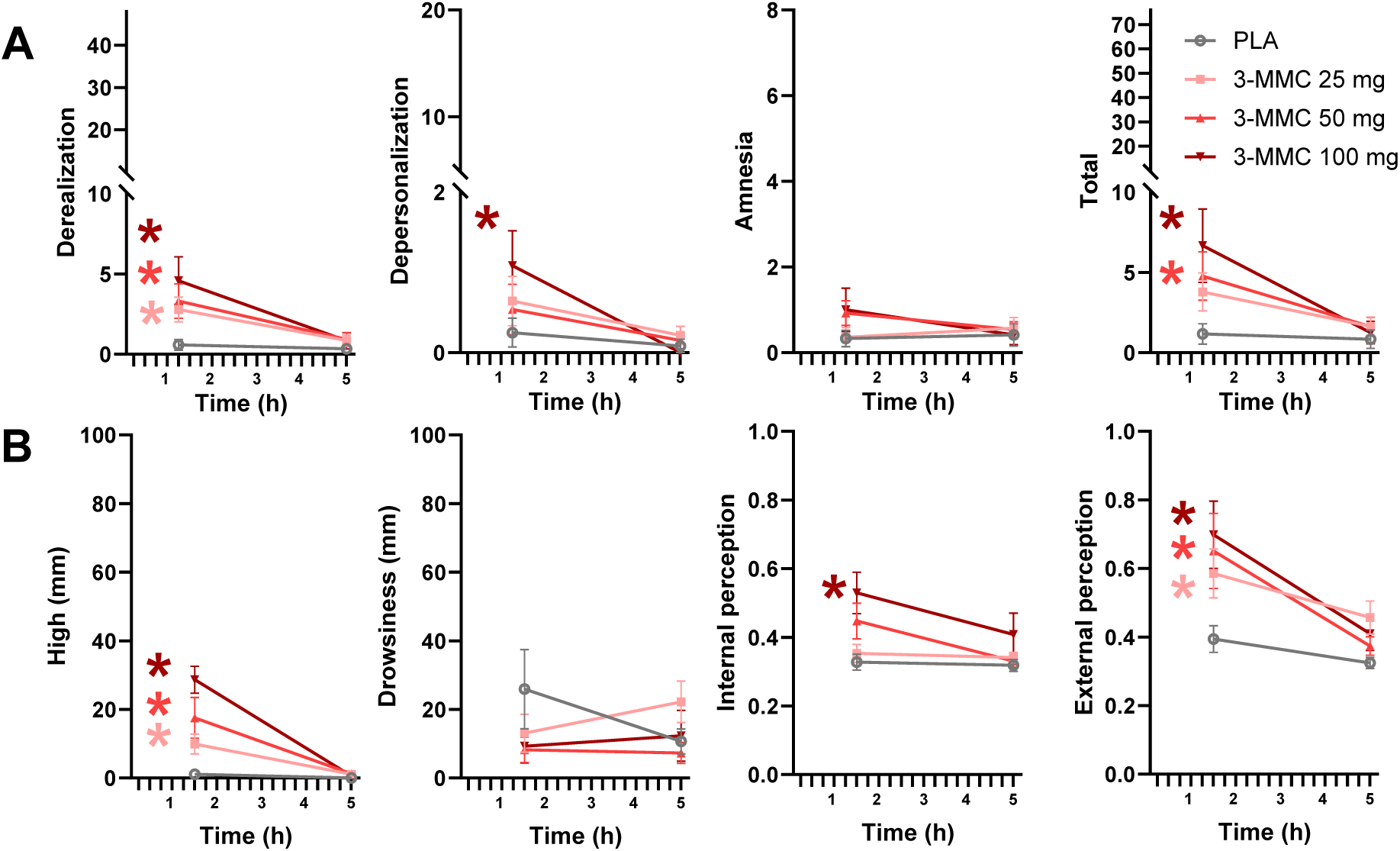
Mean (SE) subjective ratings of dissociative (panel A) and psychedelic (panel B) as assessed with the CADSS and Bowdle respectively. A * denotes a significant (p < .05) contrast between 3-MMC and placebo across all time points, with the color of the * representing the 3-MMC dose level.

Figure 4 shows subjective ratings of hunger and appetite, as well as ratings of 3-MMC wanting and liking in every treatment condition. LMM analyses showed main effects of Dose and Dose by Time on 3-MMC liking (F_3,59.813_=4.391, p=.007; F _6, 106.446_=2.786, p=.015) and wanting (F_3,65.114_=5.030, p=.003, F_6,106.630_=3.569, p=.003). Dose effects on hunger (F_3,55.753_=2.450, p=.073) and appetite (F_3, 75.681_=2.582, p=.060) failed to reach significance. Drug-placebo contrasts revealed that a dose of 100mg 3-MMC decreased appetite (p=.011), and increased ratings of drug liking (p=.007) and wanting (p=.010).

**Figure 4.**
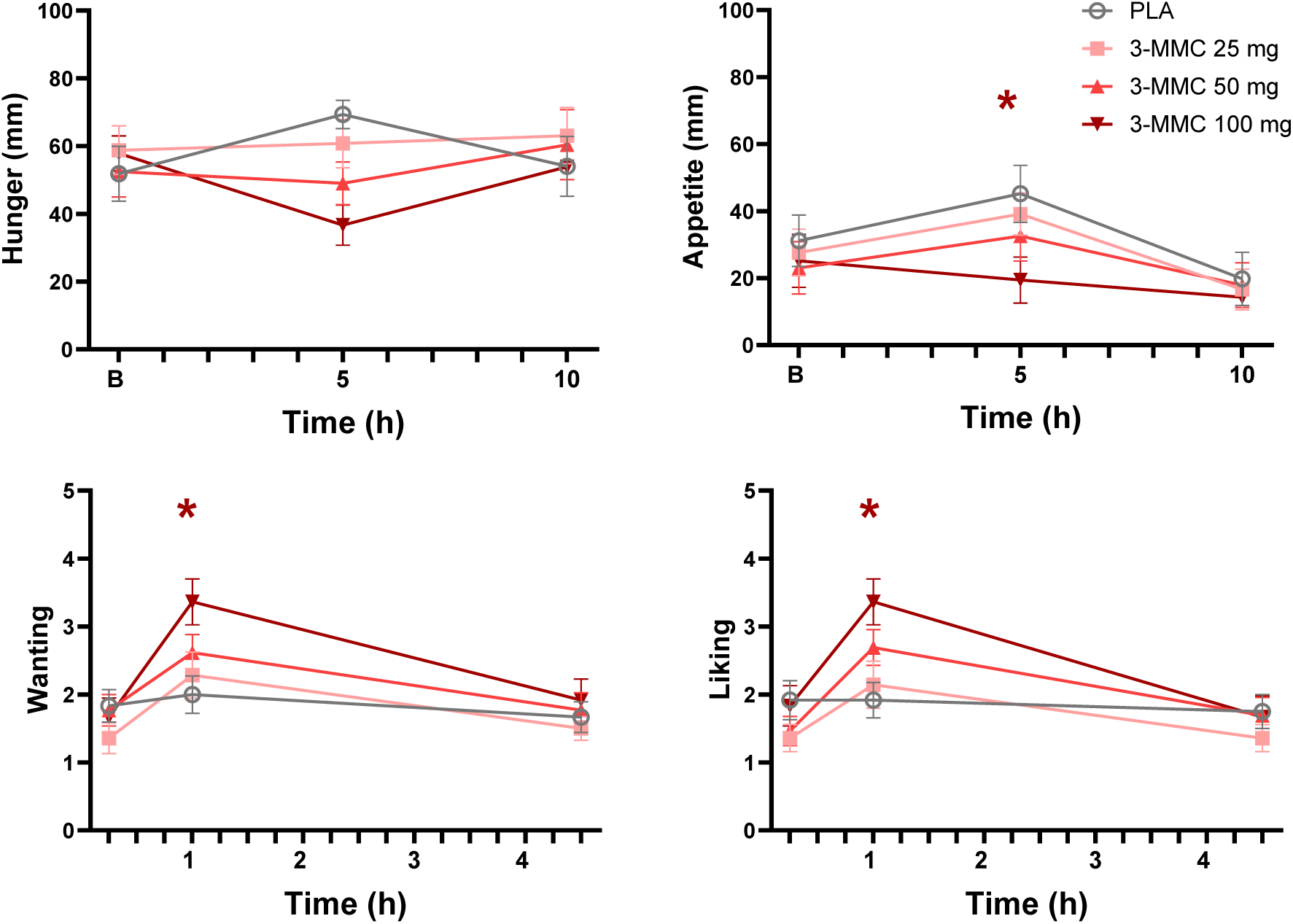
Mean (SE) subjective ratings of hunger and appetite, as well as wanting and liking of 3-MMC as a function of time after dosing. A * denotes a significant (p < .05) contrast between 3-MMC and placebo across all time points, with the color of the * representing the 3-MMC dose level.

### Adverse events

During treatment with 25mg 3-MMC there were reports of ringing in ears (1x), fatigue (1x), forgetfulness (1x) and headache (1x). Following treatment with 50mg 3-MMC reports of headache (1x) and visual flicker (1x) were noted. After treatment with 100mg 3-MMC fatigue (1x) and bruxism (1x) were reported. All adverse events were transient and mild in severity. No adverse events were reported during placebo.

## Discussion

The present study aimed to evaluate safety and cognitive pharmacodynamics of healthy volunteers after receiving single doses of 25, 50 and 100mg 3-MMC. Overall, 3-MMC produced significant, dose-related elevations in heart rate, blood pressure and subjective high. In addition, 3-MMC induced dose-related performance improvements in a range of neurocognitive domains, including processing speed, cognitive flexibility, psychomotor function, attention and memory. Subjectively, 3-MMC induced mild increments in dissociative and psychedelic effects, reduced appetite and increased ratings of 3-MMC liking and wanting.

Increments in blood pressure and heart rate are very common with stimulant drugs and usually arise from stimulation of the autonomic nervous system through the release of noradrenaline and dopamine [27,28]. In the present study, 3-MMC induced variations in blood pressure and heart rate fell largely within the normal range and did not signal any acute, adverse cardiovascular event. Cardiovascular effects were absent or mild after 25 and 50mg doses of 3-MMC and became more prominent at 100mg. The dose-relatedness of cardiovascular effects of 3-MMC suggests that their intensity might increase and potentially develop into (serious) adverse events at higher doses. Sympathomimetic effects such as agitation, tachycardia, hypertension, and hyperthermia have previously been reported for designer drugs that are structurally related to amphetamine such as mephedrone [29–31], MDMA [32], MDPV, methoxetamine, 6-ABP [33], and 4-FA [34], sometimes resulting in fatalities [30,31,35,36]. 3-MMC has also been associated with a series of intoxications in individuals who presented to hospitals with clinical manifestations of tachycardia, agitation, and hyperthermia [18]. Serum 3-MMC concentrations in these cases ranged between 2 to 1490 ng/mL (median of 91 ng/mL). Mean 3-MMC concentrations (up to 200ng/mL) as observed in the present study after low and moderate doses overlap with the lower part of this wide, clinical range. Yet, blood samples of clinical cases were not taken acutely but after treatment in the hospital or during the following day, suggesting that their residual 3-MMC concentrations resulted from the use of (very) high doses of 3-MMC at a much earlier stage. Moreover, the co-occurrence of other psychoactive substances was very common, and “mono intoxication” of 3-MMC was found in 4 out of 50 cases only [18]. A high level of 3-MMC (800 ng/mL) was also detected in a suicidal death case, from blood taken 2 days after the body was found, again suggesting the use of an extremely high dose [37]. Overall, the present study confirms the clinical notion that 3-MMC can produce cardiovascular effects, but also indicates that such effects are dose-dependent and may only become adverse and of clinical relevance at high doses. In addition, only a few adverse events were recorded after treatment with 3-MMC doses in the present study, and those that were recorded were mild and transient by nature, and of no clinical relevance. Together these findings indicate that low and moderate doses of 3-MMC were well tolerated in the present study and produced cardiovascular effects that were well within the normal range.

3-MMC enhanced task performance in a range of neurocognitive domains. Dose-related increments were observed in tracking performance and attention in the CTT and DAT, processing speed and cognitive flexibility in the DSST, spatial memory in the SMT, and performance efficiency in the MFFT. Similar improvements in neurocognitive function have been documented for various stimulant drugs, including cocaine, MDMA, amphetamine, modafinil, methylphenidate [38–47] and designer drugs such as 4-FA [34]. Psychostimulant properties of amphetamine-like compounds have long been recognized and utilized in medical applications to treat attention deficit disorders [48,49], and in military applications to maintain and sustain vigilance during operations [50,51]. It should be noted that enhancement of neurocognitive function is usually observed after low doses of psychostimulants and that the use of high doses can produce effects of overstimulation that are well outside the range of cognitive enhancement such as agitation, confusion, disturbed thinking, and even psychosis [49]. Psychostimulant properties of 3-MMC are likely caused by an increase in availability of dopamine and noradrenaline through blockage of the dopamine and noradrenaline transporters [9] as also observed with other stimulants such as amphetamine, methylphenidate, mephedrone and bupropion that share the same pharmacological mechanism [52–54]. It is noteworthy in this context that presumed increments in dopamine and noradrenaline after 3-MMC did not (negatively) affect cognitive and motor impulsivity in the present study, a finding that was previously also noted with other stimulants such as amphetamine, MDMA, methylphenidate, bupropion and 4-FA [52,55–59]. It is unclear at this point whether increments in impulsive behaviors would become prominent with higher doses of 3-MMC. Overall, the present study indicates that low to moderate doses of 3-MMC produce a profile of performance enhancement similar to other stimulants.

3-MMC produced small but consistent changes in the subjective experience of consciousness as assessed with the CADSS and Bowdle scale. Symptoms of derealization, depersonalization, subjective high, internal and external perception increased in a dose-dependent manner at the time that 3-MMC reached peak concentrations (1.5 hours after ingestion, c.f. Figure 1). The intensity of these experiences however was very low and comparable to those reported with moderate doses of other stimulants such MDMA, cocaine and 4-FA [60–63], but negligible in comparison to effects observed after a potent dissociative like ketamine, which were 4-5 times as high [63,64]. This is not to say that intense dissociative or psychedelic effects may never occur with 3-MMC, as amphetamine abuse of high doses has been associated with disturbed thinking and symptoms of psychosis in clinical cases [49,65]. Hallucinations and states of confusion have indeed been reported in cases of 3-MMC abuse [12,66]. However, the present study suggests that single administrations of low and moderate doses of 3-MMC do not seem to hold a propensity to induce strong dissociative or psychedelic effects.

Self-ratings of drug liking and wanting, considered sensitive indicators of the potential for drug abuse, were elevated under 3-MMC as compared to placebo. Ratings of drug liking and wanting were most prominent during peak concentrations of 100 mg 3-MMC, but returned to baseline at 5 hours after administration. This appears consistent in view of the rather short elimination half-life of about 3 hours. Likewise, ratings of appetite after lunch decreased after 100mg of 3-MMC as compared to placebo, but these also returned to baseline at 10 hours after administration. These findings suggest that low to moderate doses of 3-MMC are unlikely to lead to sustained feelings of drug liking and wanting or sustained appetite suppression that may lead to repeated or compulsive use, however, it cannot be excluded that the short-lived effects may trigger repeated ingestion during short periods. Yet again, abuse liability of 3-MMC, as with any amphetamine [49], may increase with the use of higher doses. Abuse liability after (repeated) use of high doses of 3-MMC has not been systematically studied in humans but preclinical evidence has suggested addictive properties of 3-MMC [67] and clinical cases of 3-MMC dependence have been reported [68].

The present study is the first to assess the safety profile of the designer drug 3-MMC. Where new pharmaceutical drugs must undergo rigorous rounds of safety checks in extensive clinical trials before they can be approved for human use, this is typically not the case for designer drugs that come from clandestine chemistry labs. Hence, potential harms caused by designer drugs that enter the consumer market are basically unknown. The general lack of safety data on designer drugs also hampers accurate risk evaluations by public health organizations that are mostly limited to basic pharmacology, anecdotal clinical case reports and expert opinion. Full-scale evaluations of health risks associated with designer drugs are virtually missing because there is a general lack of controlled studies with these compounds, often due to ethical and legal constraints. The present evaluation of the risk profile of low and moderate doses of 3-MMC therefore complements and nuances existing risk evaluation [4,5] that are solely based on extreme cases of fatal and non-fatal poisonings in which 3-MMC, mostly in combination with other drugs, was implicated. Though important, extreme cases of 3-MMC abuse are not representative of risks that may be endured among the majority of 3-MMC users who consume the drug recreationally and avoid excessive use. The current study suggests that in this type of user, the safety risks of 3-MMC at single doses up to 100 mg are very low to negligible. This is relevant as also public health committees have noted the importance of collecting additional and accurate knowledge on 3-MMC that can be disseminated to people who use 3-MMC, as well as to practitioners, policymakers and decision-makers [5].

In summary, low to moderate doses of 3-MMC produced cardiovascular effects that remained within the normal range and improved performance across a range of neurocognitive domains in a dose-related manner. 3-MMC induced mild dissociative and psychedelic effects and transiently increased ratings of 3-MMC liking and wanting. Overall, the cardiovascular, psychostimulant and psychotomimetic profile of 3-MMC appears in line with that of compounds that are structurally related to amphetamine. It is concluded that low to moderate doses of 3-MMC were well tolerated and safe, and that potential health risks might only occur at high or excessive doses of 3-MMC.

## Data Availability

All data produced in the present study are available upon reasonable request to the authors

## Author contributions

Design and conceptualization – JGR, ELT; Data acquisition – JTR, NLM; PK analysis – SWT, Statistics and data analysis – JGR; Writing – JGR; Editing and review of the manuscript - all.

## Acknowledgements

The authors thank the members of the SSG (Cees van Leeuwen, Tomas Palenicek, Rudy Schreiber) for evaluating the safety data and providing advice.

